# Assessing Early-Life Contributions to Racial Disparities in Cognition for Older Americans: The Importance of Educational Quality and Experience

**DOI:** 10.1101/2023.10.05.23296614

**Authors:** Zhuoer Lin, Justin Ye, Heather Allore, Thomas M. Gill, Xi Chen

**Author notes:** Correspondence to: Xi Chen, Department of Health Policy and Management, Yale School of Public Health, 60 College St, New Haven, CT 06520, United States of America. Joint first authors with equal contribution (Zhuoer Lin, Justin Ye).

## Abstract

**Importance:** Studies on racial disparities in cognition have primarily focused on mid-life to late-life risk factors. Given the critical role of neurocognitive development in early life, understanding contributions of early-life circumstances has important implications for preventive strategies.

**Objective:** To assess the extent to which racial differences in early-life circumstances, particularly educational quality and experience, can individually and collectively contribute to racial disparities in late-life cognition.

**Design, Setting, and Participants:** 9,015 participants (7,381 non-Hispanic White (White) and 1,634 non-Hispanic Black (Black) with comprehensive life-history data were assembled from the Health and Retirement Study (HRS) (1995-2018), a nationally representative longitudinal survey of Americans 50 years or older.

**Main Outcomes and Measures:** Blinder-Oaxaca Decomposition (BOD) was used to quantify disparities in cognitive outcomes between White and Black participants attributable to racial differences in early-life circumstances. Cognitive score and cognitive impairment were assessed using the Telephone Interview for Cognitive Status (TICS). Early-life educational quality and experience were included as key explanatory variables, and traditional early-life factors including cohort, regional, financial, health, trauma, relationship factors, and educational attainment were included as additional explanatory variables. Demographic and genetic factors were included as covariates.

**Results:** Among White and Black participants, the mean (SD) ages were 73.2 (10.1) and 69.2 (9.2) years, respectively, and 4,410 (59.7%) and 1,094 (67.0%) were female. Cognitive scores (range: 0-27 points) were significantly lower in Black (13.5, 95% CI, 13.3-13.7) than White participants (15.8, 95% CI, 15.7-15.9), while the prevalence of cognitive impairment was significantly higher in Black (33.6, 95% CI, 31.3-35.9) than White participants (16.4, 95% CI, 15.6-17.2). Substantial racial differences were observed in early-life circumstances. Overall, racial differences in early-life circumstances accounted for 65.9% and 85.1% of the gaps in cognitive score and impairment, respectively. Educational quality and experience played a prominent role, independently explaining 34.1% of the gap in cognitive score and 51.3% in cognitive impairment. Notably, school racial segregation (attending all minority schools before college) explained 26.7%-40.6% of the gaps in cognition. These findings remained consistent after adjusting for genetic risks.

**Conclusions and Relevance:** Less favorable early-life circumstances contribute to clinically meaningful and statistically significant racial gaps in late-life cognition. Policies that improve educational equity may have long-lasting impacts on reducing racial disparities in cognition into older ages.

**Key Points:** *Questions:* How much do differences in early-life circumstances, including educational quality and experience, individually and collectively explain late-life disparities in cognitive outcomes between non-Hispanic Black (Black) and non-Hispanic White (White) older adults?

*Findings:* Early-life circumstances contribute substantially to racial disparities in cognitive outcomes over age 50. Educational quality and experience are the most important early-life contributors, independent of a rich set of sociodemographic and genetic factors.

*Meaning:* Exposure to less favorable early-life circumstances for Black than White adults is associated with large racial gaps in cognitive outcomes.

## Introduction

There are marked differences in rates of cognitive impairment, including dementia, among the growing US population of older adults. The prevalence of dementia for non-Hispanic Black (Black) adults is two to three times that of non-Hispanic White (White) adults, after accounting for age, sex, education, and late-life comorbidities.^1–3^ The rising proportion of US older adults who are minorities may lead to a considerable rise in socioeconomic burden associated with cognitive impairment and dementia,^4–7^ of which growing evidence suggests that low education, physical inactivity, smoking, and social isolation are among the strongest risk factors. ^8,9^ Most research has focused on mid-life to late-life factors, while the early-life circumstances through which the racial gap in cognition may arise have received much less attention.

There is a lack of evidence linking factors contributing to neurocognitive development earlier in life with cognitive impairment and dementia in later life. However, brain development is most rapid and plastic early in life.^10,11^ Strong early-life brain development supports more complex neuritic and intraneuronal connections and cognition, conferring a young adulthood and middle age advantage^10^ that may be associated with a more robust cognitive reserve and a lower risk of dementia in later life.^12,13^ Early life education represents a particularly important modifiable risk factor.^8^ In addition to years of education, quality and experience of education may affect brain health. For instance, exposure to a higher concentration of minority students and segregated schools is associated with diminished occupational aspirations, expectations, and achievement among minority students.^14,15^ The possible long-term health consequences of educational factors and their implications for racial inequities are less well known, especially when accounting for various other early-life socio-environmental exposures.^16–19^

Using a nationally representative longitudinal survey with up to 24 years of follow-up and a unique set of life history data, we investigated whether and how racial gaps in cognitive status and prevalence of cognitive impairment among older Americans are tied to racial differences in early-life circumstances. We tested the following hypotheses: 1) early-life circumstances contribute significantly and sizably to racial disparities in cognitive outcomes; 2) educational quality and experience is the most important early-life contributor to racial cognitive gaps in old age, independent of years of education, other demographic and other factors.

## Methods

### Study Design and Participants

We used the Health and Retirement Study (HRS), a large nationally-representative, longitudinal study of older Americans (aged ≥ 50 years). We assembled a large array of factors on early-life circumstances from three HRS components: the core survey (1995-2018); the Life History Mail Survey (LHMS) (2015, 2017); the Enhanced Face-to-Face (EFTF) Interview (2006-2012); In addition, we obtained polygenic risk scores (PGS), which were estimated from the saliva samples in the EFTF interviews (2006-2012). Details of these data sources are provided in eAppendix A and elsewhere.^20^ The study followed the Strengthening the Reporting of Observational Studies in Epidemiology (STROBE) guideline.

Our analysis included HRS LHMS participants who had at least one cognitive assessment from 1995 to 2018. The sample selection criteria are provided in Figure 1. We excluded participants who did not self-identify as White or Black adults and those who did not participate in EFTF. The resulting analytical sample with both LHMS and EFTF data included 9,015 participants (White: n = 7,381; Black: n = 1,634) who had data on a wide spectrum of early-life factors. Their latest wave, i.e., most proximate to 2018, of cognitive assessment and demographic covariates from the same core survey were included to optimize sample size. The 7,513 participants (6,260 White, 1,253 Black) who had valid measures of PGS were further included in the sensitivity analysis.

**Figure 1.**
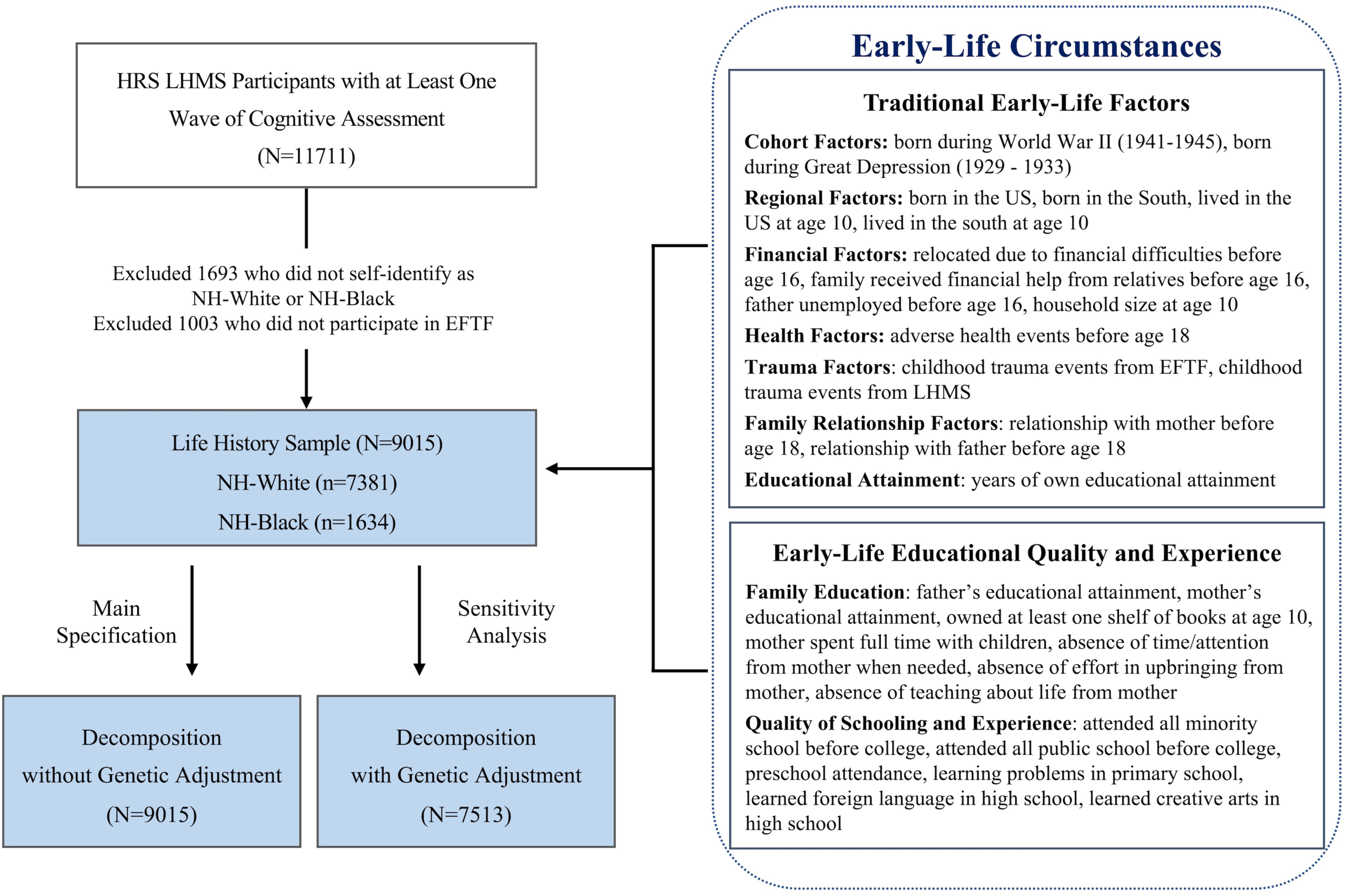
Flow Chart of Sample Selection Process. *Notes*: HRS=Health and Retirement Study; EFTF=Enhanced Face-to-Face; LHMS=Life History Mail Survey; NH=non-Hispanic. EFTF and LHMS are separate components conducted within HRS.

The HRS study protocol was approved by the Institutional Review Board at the University of Michigan. Data used in this study are de-identified and publicly available.

### Cognitive Outcomes

The HRS assessed cognitive function with a range of tests adapted from the Telephone Interview for Cognitive Status (TICS), which has high validity among White and Black adults.^21,22^ Specifically, the 27-point cognition scale represents three cognitive tests: an immediate and delayed 10-noun free recall test to measure memory (0 to 20 points); a serial sevens subtraction test to measure working memory (0 to 5 points); and a counting backwards test to measure speed of mental processing (0 to 2 points). Built on existing criteria, in each wave of cognitive tests, two categories were derived from the total summary score for cognition (0 to 27 points): normal (12 to 27 points) and cognitively impaired (0 to 11 points).^22,23^ The latter group included participants ranging from mild cognitive impairment to dementia. Dementia was not separately analyzed due to small numbers.^24^ Nineteen participants with only proxy cognitive responses (<0.2%) were not included.

### Early-Life Educational Quality and Experience

Our analysis incorporated a comprehensive set of measures related to early-life educational quality and experience as key explanatory variables, which were uniquely collected in the HRS LHMS. These measures consist of two important components: family education and quality of schooling and experience (Figure 1). To assess family education, we included the number of years of parental education, whether the respondent owned at least one shelf of books at age 10, whether the mother spent full time with children, whether she was absent in terms of time/attention, her effort in upbringing, and her teaching before the respondent turned 18. To evaluate the quality of schooling and experience, we considered whether the respondent attended all minority (segregated) schools before college, attended all public schools before college, attended preschool, had learning problems in primary school, learned a foreign language in high school, and learned creative arts in high school. A more detailed description of variable definition and construction is presented in eAppendix B.

### Traditional Early-Life Factors

Our study integrated a wide spectrum of early-life circumstances that were selected following a thorough review of the existing literature, spanning seven key domains that encompassed early-life cohort, regional, financial, health, trauma, family relationship factors, and educational attainment. They are referred to hereafter as traditional early-life factors, and the factors are itemized in Figure 1. A detailed description of these early-life circumstances can be found in eAppendix B.

### Covariates

Demographics (i.e., age, sex, and marital status) and genetic factors were included as covariates. Genetic factors were assessed through the use of PGS (eAppendix A & B), enabling the inclusion of biological predispositions associated with anthropometrics, diseases, cognition and longevity, mental health and personality, and smoking.^25,26^

### Statistical Analysis

To compare the characteristics between Black and White participants, we used Welch t-tests for continuous variables, Mann-Whitney-Wilcoxon tests for ordinal variables, and Chi-square tests for categorical variables.^27^ Additionally, for each of the early-life circumstances, we estimated the standardized differences between Black and White participants.

To evaluate racial disparities in cognitive outcomes, we performed a Blinder-Oaxaca Decomposition (BOD) to attribute these disparities to differences in early-life circumstances *versus* differences in the effects of these circumstances.^28^ All decompositions were formulated based on Black participants, and the contribution of early-life circumstances was characterized as the expected change in cognitive outcomes of Black participants if they had the same mean predictor values as White participants.^29^ A more detailed description of the BOD and estimation procedures can be found in eAppendix C.

For each cognitive outcome, we conducted two decomposition models: LifeHistory**_1_** (Model 1), which included traditional early-life factors and other covariates; and LifeHistory**_2_** (Model 2), which also included early-life educational quality and experience. Each decomposition assessed the contribution of factors at both the overall level (i.e., the racial gap in cognitive outcomes explained by all early-life circumstances combined) and the factor/domain level (i.e., the racial gap to which each individual factor/domain of circumstance independently contributed, adjusted for the others). Genetic factors were included as covariates only in the sensitivity analysis due to diminished sample size (as shown in Figure 1).

To address item-level missingness of early-life factors and covariates, we performed multiple imputation,^30^ following procedures adopted in the HRS. We used a sequential regression approach (i.e., chained equations) that creates imputations through a sequence of multiple regressions.^31,32^ The imputation models included all the early-life factors (except genetic factors), covariates in the decomposition analyses, and other non-changing baseline demographics (e.g., birth year).^31,32^ Following existing guidelines, 20 imputed datasets were produced and analyzed; and the parameter estimates for each imputed dataset were pooled/combined to obtain the final decomposition results.^33,34^ Genetic factors were not imputed, and all participants included in the analyses had valid measurements of cognitive outcomes. Additional details about variable missingness and imputation of early-life factors and covariates are provided in eAppendix B.

The analyses were carried out using STATA (version 17.0), IVEWare (version 0.3) and R (version 4.0.2). All tests were two-sided with an alpha level of 0.05 for statistical significance.

## Results

### Characteristics of the Study Population

Table 1 shows descriptive statistics for cognitive outcomes, covariates, and early-life circumstances of the study population. Compared to White participants, Black participants had lower cognitive scores and higher proportion of cognitive impairment. In the Life History sample, the differences in cognitive scores between White and Black participants were 2.3 (95% CI, 2.1-2.6; *P*<.001) points for cognitive score, and 17.2 (95% CI, 14.8-19.6; *P*<.001) percentage points (pp) for cognitive impairment. Additionally, Black participants were on average younger than White participants, with an average gap of 4.0 (95% CI, 3.5-4.5; *P*<.001) years.

**Table 1.** Characteristics of Study Participants from the Health and Retirement Study Cohort Collected Between 1995 and 2018^a^.

|  | **Life History Sample <sup>b</sup>**<br>**(N=9015)** |  |
| --- | --- | --- |
| **Characteristic** | **Black** | **White** |
| No. of participants | 1634 | 7381 |
| **Outcomes and Demographic Covariates** |  |  |
| Cognitive score, mean (SD) | 13.5 (4.8) | 15.8 (4.4) |
| Cognitive impairment prevalence, No. (%) | 549 (33.6) | 1210 (16.4) |
| Age, mean (SD) | 69.2 (9.2) | 73.2 (10.1) |
| Female, No. (%) | 1094 (67.0) | 4410 (59.7) |
| Married/cohabiting, No. (%) | 714 (43.8) | 4666 (63.3) |
| **Traditional Early-Life Factors** |  |  |
| Born during World War II (1941-1945), No. (%) | 199 (12.2) | 1110 (15.0) |
| Born during the Great Depression (1929-1933), No. (%) | 90 (5.5) | 702 (9.5) |
| Born in the South, No. (%) | 1087 (66.6) | 1850 (25.1) |
| Born outside of the US, No. (%) | 89 (5.5) | 294 (4.0) |
| Lived in the South at age 10, No. (%) | 974 (59.6) | 2051 (27.8) |
| Lived outside of the US at age 10, No. (%) | 72 (4.4) | 196 (2.7) |
| Relocated due to financial difficulties before age 16, No. (%) | 292 (18.0) | 1147 (15.6) |
| Family received financial help before age 16, No. (%) | 347 (21.9) | 1032 (14.2) |
| Father unemployed before respondent's age 16, No. (%) | 276 (20.8) | 1465 (21.0) |
| Household size at age 10, median (IQR) | 6 (3) | 5 (2) |
| Adverse childhood health events, No. (%) | 1167 (72.1) | 6087 (83.5) |
| Childhood trauma events (EFTF), No. (%) | 557 (34.5) | 2515 (34.1) |
| Childhood trauma events (LHMS), No. (%) | 908 (56.4) | 2433 (33.4) |
| Good relationship with mother before age 18, No. (%) | 1203 (80.9) | 5538 (78.2) |
| Good relationship with father before age 18, No. (%) | 956 (69.1) | 4975 (71.9) |
| Years of educational attainment, median (IQR) | 12 (2) | 13 (4) |
| **Early-Life Educational Quality and Experience** |  |  |
| Years of father's educational attainment, median (IQR) | 8 (6) | 12 (4) |
| Years of mother's educational attainment, median (IQR) | 10 (4) | 12 (4) |
| Owned at least one shelf of books at age 10, No. (%) | 863 (54.1) | 5038 (69.7) |
| Mother spent full time with children before respondent's age 18, No. (%) | 349 (22.3) | 3251 (44.7) |
| Absence of time/attention from mother when needed, No. (%) | 58 (3.7) | 219 (3.0) |
| Absence of effort in upbringing from mother, No. (%) | 55 (3.6) | 179 (2.5) |
| Absence of teaching about life from mother, No. (%) | 95 (6.1) | 427 (6.0) |
| Attended all minority school before college, No. (%) | 1011 (73.5) | 44 (0.6) |
| Attended all public school before college, No. (%) | 1294 (89.6) | 5337 (76.9) |
| Preschool attendance, No. (%) | 290 (18.5) | 894 (12.4) |
| Learning problems in primary school, No. (%) | 248 (15.4) | 840 (11.5) |
| Learned foreign language in high school, No. (%) | 662 (42.5) | 3656 (51.3) |
| Learned creative arts in high school, No. (%) | 950 (60.4) | 4507 (63.2) |

Abbreviations: SD, standard deviation; IQR, interquartile range; LHMS, Life History Mail Survey (2015, 2017); EFTF, Enhanced Face-to-Face (2006-2012).
<sup>a</sup> The descriptive statistics presented in the table were obtained using non-missing data before imputation. The sample size for each variable might be lower than the total sample size, though the difference was minimal (see eAppendix B for details).
<sup>b</sup> Life History sample refers to participants who had valid cognitive measure and participated in the LH

Black and White participants had many differences in early-life circumstances (Figure 2). In terms of cohort and regional factors, Black participants were more likely born in the South (*P*<.001) and living in the South at age 10 (*P*<.001) than White participants. They also had less favorable early-life socioenvironmental factors than White participants. Notably, they were more likely to relocate due to financial difficulties (*P*=.016) and receive financial help (*P*<.001) during childhood than White participants. Moreover, they had a larger household size at age 10 (*P*<.001) and were more likely to experience early-life trauma (*P*<.001 for LHMS items) as compared to White participants.

**Figure 2.**
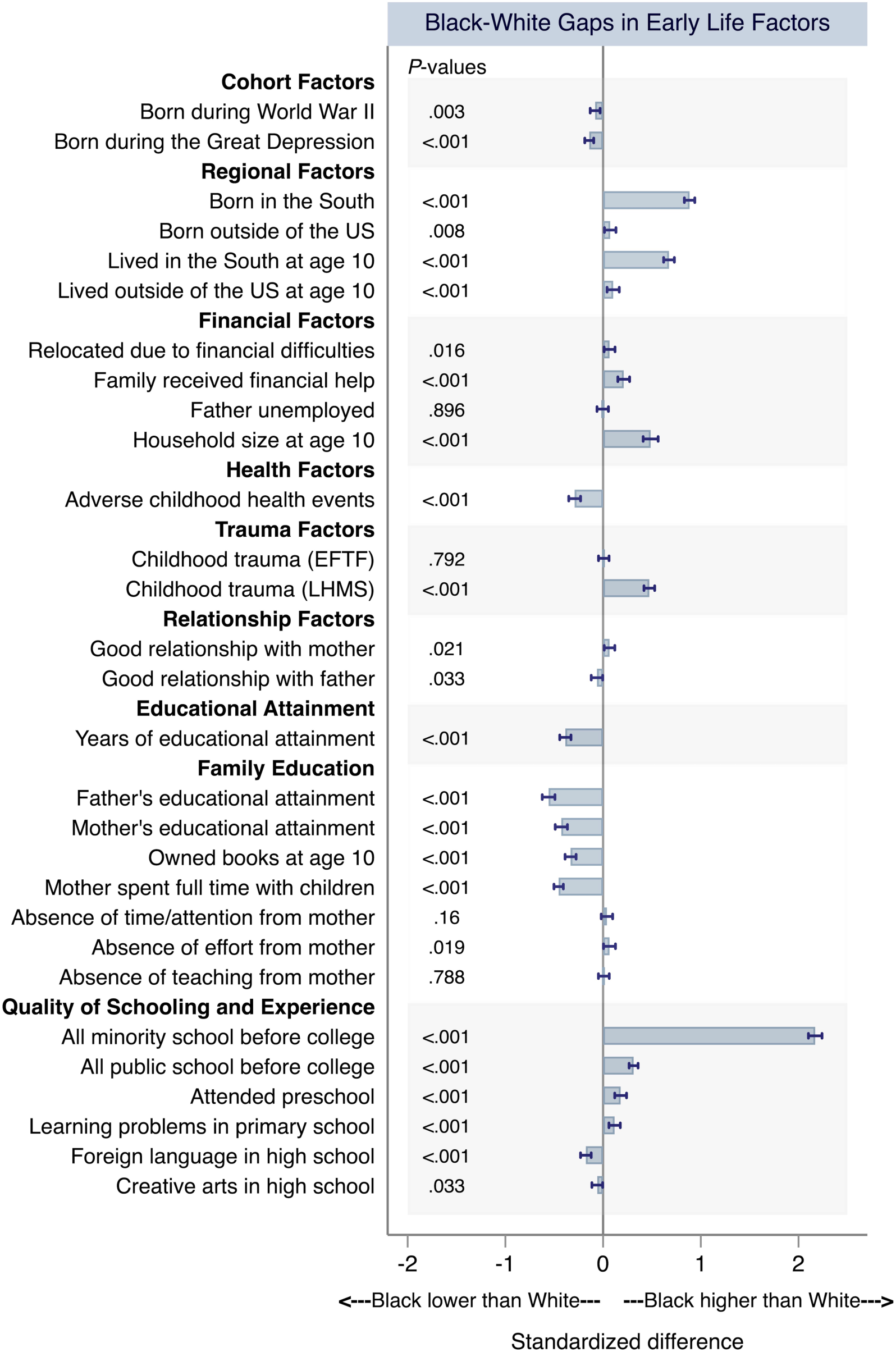
Standardized Differences in Early-Life Circumstances between Black and White Participants from the Health and Retirement Study Cohort Collected between 1995 and 2018 (N=9015) ***Notes*:** The figure presents the standardized differences in individual early-life circumstances between Black and White participants. For each early-life factors, the mean difference between Black and White participants was standardized by its standard deviation; and the estimates were visualized in the figure. The horizontal bar denotes the standardized differences between Black and White participants for each individual early-life factors, and the horizontal line denotes the 95% confidence interval. The p-values listed alongside the y axis denote the statistical significance of the differences. A positive (negative) value indicates Black participants had higher (lower) value of early-life factor than White participants. The differences were estimated using Life History sample without data imputation and sample size for each factor can be slightly less than 9,015 due to item-level missingness (see eAppendix B for more details). The results were similar when using imputed data or adjusting for sample demographics (available upon request).

Stark racial differences were observed in educational attainment and educational quality and experience. Black participants had significantly lower educational attainment (*P*<.001) and lower parental educational attainment (*P*<.001) than White participants. A lower proportion of Black participants owned books at age 10 (*P*<.001), and their mothers were less likely to spend full time with them during childhood (*P*<.001). The quality of schooling and experience were also less advantaged among Black participants. Specifically, relative to White participants, Black participants were more likely to report learning problems in primary school (*P*<.001), a lower proportion learned a foreign language in high school (*P*<.001), and much higher proportions attended all minority schools (*P*<.001), and public schools before college (*P*<.001).

### Decomposing Overall Early-life Contributions

Figure 3 presents the overall contributions of differences in early-life circumstances to disparities in cognitive score and cognitive impairment between White and Black participants (see eTables 1-2 for the numerical estimates).

**Figure 3.**
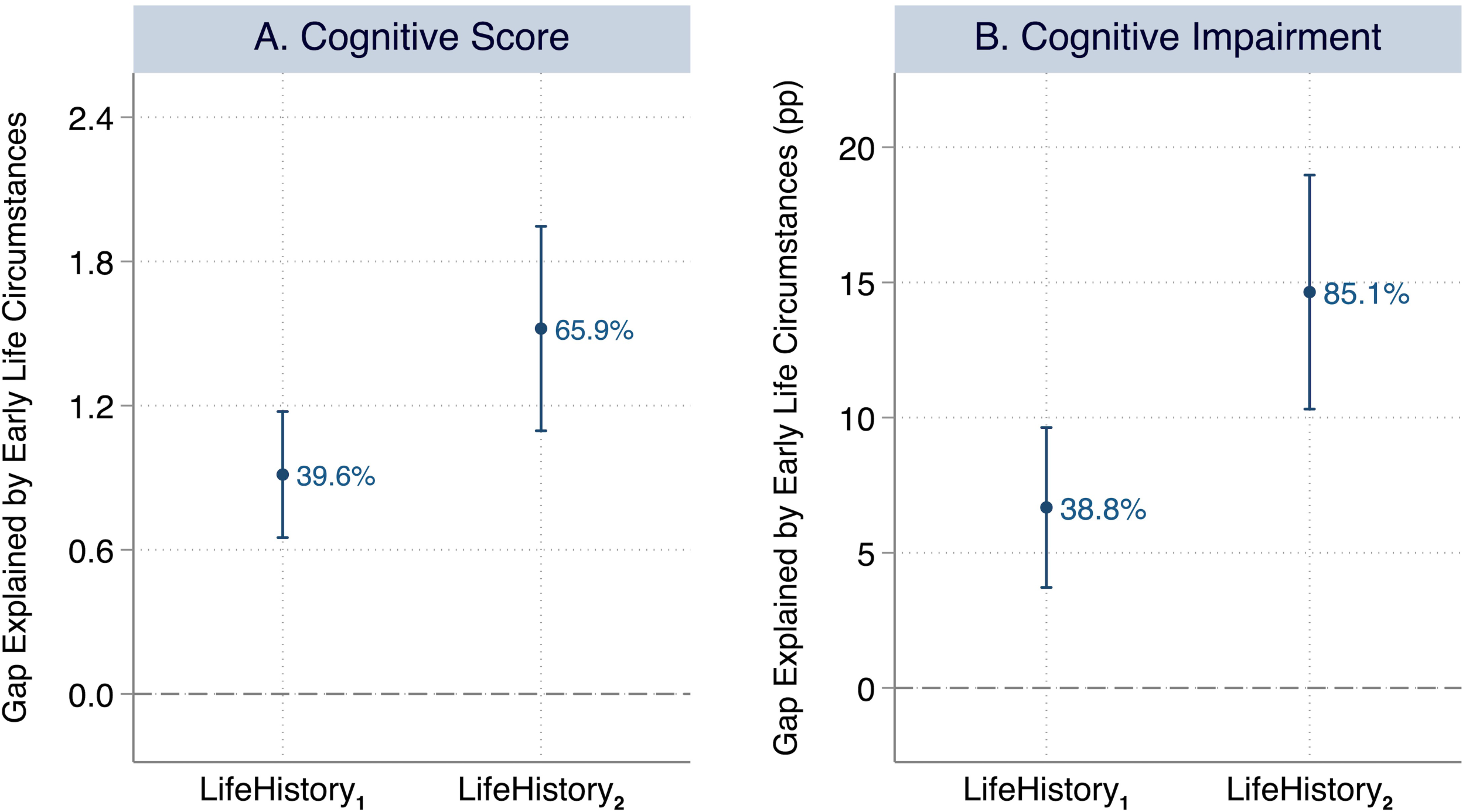
Absolute and Relative Contribution of Overall Early-Life Circumstances to Racial Gaps in Cognition between White and Black Participants (N=9015) ***Notes*:** Panel A presents the decomposition results for cognitive score, a continuous variable ranging from 0 to 27; and Panel B presents the decomposition results for cognitive impairment, a dichotomous variable indicating whether individuals were classified as cognitive impaired (0/1). For each cognitive outcome, X axis denotes the models being examined, including LifeHistory_1_, LifeHistory_2_. In LifeHistory_1_, traditional early-life factors were included to perform the decomposition. In LifeHistory_2_, early-life educational quality and experience were also included. All decompositions adjust for demographic covariates including age, sex and marital status. In Panels A and B, Y axis denotes the racial gap in cognitive outcomes between White and Black participants that was explained by their differences in early-life circumstances, representing the absolute contribution of early-life circumstances to racial gaps in cognition. In Panel A, the unit of cognitive score is point; and in Panel B, the unit of cognitive impairment is percentage point (pp). The point estimates are plotted as circles and their 95% confidence interval are plotted as vertical lines. The relative contributions of the early-life circumstances (%) in each setting are also presented alongside the vertical lines, denoting the percentage of gaps in cognition that was explained by early-life circumstances. The pooled estimates were obtained using multiple imputation with 20 imputed datasets.

Differences in early-life circumstances accounted for 0.91 (95% CI, 0.65-1.2) points, or 39.6% of the racial gap in cognitive score between White and Black participants in analyses that included only traditional early-life factors (LifeHistory**_1_**) and demographic covariates. The contribution substantially increased when educational quality and experience were also included, with differences in early-life circumstances accounting for 1.5 (95% CI, 1.1-2.0) points or 65.9% of the racial gap in cognitive score (LifeHistory**_2_**).

The results for cognitive impairment were generally comparable, although the contribution of early-life circumstances was larger than that for cognitive score in LifeHistory_2_. Across LifeHistory_1_ and LifeHistory_2_, differences in early-life circumstances explained 6.7 (95% CI, 3.7-9.6) pp or 38.8%, and 14.6 (95% CI, 10.3-19.0) pp or 85.1% of the racial gaps in cognitive impairment, respectively.

The contribution of early-life circumstances did not change substantively after the addition of genetic factors as covariates (eFigure 2).

### Deciphering Main Contributors in Early Life

Figure 4 and eTable 2 show the factor- and domain-level contributions of early-life circumstances in LifeHistory**_2_**.

**Figure 4.**
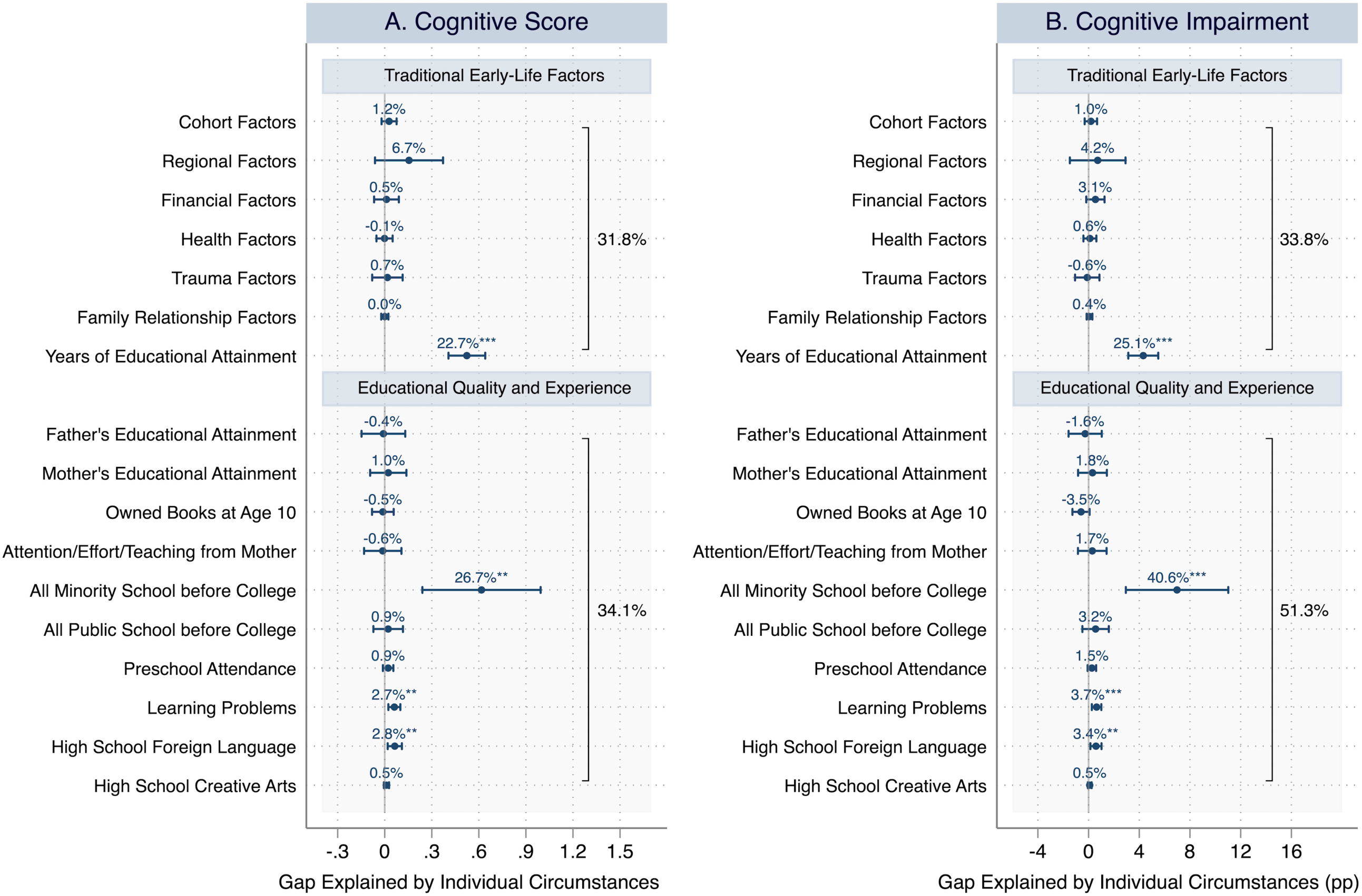
Absolute and Relative Contribution of Individual Early-Life Circumstances to Racial Gaps in Cognition between White and Black Participants, LifeHistory**_2_** (N=9015) ***Notes*:** Panel A shows the decomposition results for cognitive score, a continuous variable ranging from 0 to 27; Panel B shows the decomposition results for cognitive impairment, a dichotomous variable indicating whether individuals were classified as cognitive impaired (0/1), or not. For each cognitive outcome, Y axis denotes the variables or domains being examined; X axis denotes the amount of racial gap in cognition between White and Black participants that was explained by their differences in the individual early-life circumstances (i.e., absolute contribution). In Panel A, the unit of cognitive score is point; and in Panel B, the unit of cognitive impairment is percentage point (pp). The point estimates are plotted as circles and their 95% confidence interval are plotted as horizontal lines. The relative contributions of the early-life circumstances (%) in each setting are presented alongside the horizontal lines, denoting the percentage of gaps in cognition that was explained by early-life circumstances. The decompositions were performed using Life History sample without genetic adjustment (LifeHistory**_2_**), and the numerical results can be found in Supplementary eTable 2. The pooled estimates were obtained using multiple imputation with 20 imputed datasets. Asterisks denote the statistical significance of the contribution: *** *P* < 0.001, ** *P* < 0.01, * *P* < 0.05.

Among all early-life factors, differences in educational quality and experience contributed most substantively to the racial gaps in cognition, accounting for 0.79 (95% CI, 0.38-1.19) points or 34.1% in cognitive score and 8.8 (95% CI, 4.6-13.1) pp or 51.3% in cognitive impairment, independent of traditional early-life factors and covariates. Collectively, differences in traditional early-life factors contributed slightly less to the racial gaps in cognition, accounting for 0.73 (95% CI, 0.46-1.0) points or 31.8% in cognitive score, and 5.8 (95% CI, 3.2-8.4) pp or 33.8% in cognitive impairment.

For educational quality and experience, participants who attended minority schools had significantly lower cognitive score and higher proportion of cognitive impairment, especially among Black participants (eFigure 1). Attending all minority school before college contributed the most to the racial disparities, accounting for 0.62 (95% CI, 0.24-0.99) points or 26.7% of the racial gap in cognitive score and 7.0 (95% CI, 2.9-11.0) pp or 40.6% of the gap in cognitive impairment between White and Black participants. Among the other factors of educational quality and experience, learning problems in primary school and not learning a foreign language in high school, respectively, accounted for 2.7% and 2.8% of the racial gap in cognitive score, and 3.7% and 3.4% of the gap in cognitive impairment.

Among the traditional early-life factors, years of educational attainment contributed the most to racial gaps in cognitive outcomes, accounting for 0.52 (95% CI, 0.41-0.64) points or 22.7% of the racial gap in cognitive score, and 4.3 (95% CI, 3.1-5.5) pp or 25.1% of the racial gap in cognitive impairment between White and Black participants. Although birth region accounted for 6.7% of racial gap in cognitive score, this contribution was not statistically significant. Cohort, financial, health, trauma, and relationship factors did not contribute substantively to the racial disparities in cognition.

These results did not change substantively after the addition of genetic factors as covariates (eFigure 3).

## Discussion

Few prior studies have evaluated associations between a rich set of early-life circumstances and racial gaps in cognitive outcomes. In this longitudinal study of older Americans, we evaluated to what extent two clinically meaningful cognitive outcomes differ between Black and White older adults, and quantified how much early-life circumstances, including educational quality and experience, contributed collectively and individually to the racial gaps. Three major findings warrant comment.

First, our study revealed substantial disparities between Black and White adults in both early-life circumstances and later-life cognition. The presence of less favorable early-life social-demographic circumstances among Black older adults, relative to their White counterparts,^35,36^ was associated with a clinically meaningful racial gap in cognitive outcomes, with Black older adults doing more poorly. Our findings underscore the substantial impact of early-life circumstances in elucidating cognitive outcome disparities among racial groups, even surpassing their explained share of overall cognitive variability.^8^ From a policy perspective, addressing adverse early-life circumstances has the potential to alleviate racial disparities in late-life cogniton.^35^

Second, among the early-life social-demographic circumstances lower educational quality and experience among Black participants were the greatest contributors to racial gaps in cognitive outcomes, even after accounting for demographic and genetic factors. Educational factors may work through a set of channels to influence racial gaps, such as brain development, employment, income, access to health care, and lifestyle choices.^37^ For example, attending minority (e.g., segregated) schools may expose Black participants to discrimination and racism that contributed to stress, traumatized brain development, and behavioral problems.^38,39^ Overall, our results suggest that policies implemented to improve equitable education may generate long-lasting impacts on reducing racial disparities in cognitive outcomes into older ages.^40,41^

Third, our findings suggest that the associations between educational quality and experience and cognitive outcomes cannot be solely attributed to educational attainment. This underscores that relying solely on educational attainment to gauge the relationship between education and cognitive impairment might be insufficient. In fact, the racial differences in educational quality and experience itself contributed a larger portion to the racial gaps in cognitive outcomes than educational attainment did.

The escalating socioeconomic disparities over the last four decades^35,42^ have led to a situation where the offspring of younger minority parents face exacerbated early-life disadvantages, potentially amplifying racial disparities in cognitive outcomes.^43–44^ Meanwhile, the heightened flows of migration in recent decades and the implementation of educational policies aimed at countering segregation have likely contributed to the desegregation of schools, consequently narrowing racial gaps.^45^ Therefore, continuously monitoring the trend in racial gaps among upcoming age cohorts and generations is warranted.

This analysis builds on prior work that has evaluated the association between education and later-life cognitive outcomes. An important strength of the current analysis is the focus on the role of early-life circumstances prior to full educational attainment. A second major strength relates to our accounting for many early-life circumstances, which enabled us to better understand how they individually and collectively shape racial disparities in late-life cognition. To our knowledge, the current study is the first to quantify the contributions made by this uniquely rich set of early-life circumstances. The third major strength of this study involves our in-depth investigation into the important but often neglected role played by educational quality and experience.

### Limitations

Our observational study has several limitations. First, persons with impaired cognition or other health problems, as well as those with more disadvantaged early-life circumstances, may have been less likely to participate, suggesting that the associations between early-life circumstances and late-life racial disparities in cognition may be conservative. Second, early-life circumstances in this study were self-reported, which may introduce recall bias. Third, due to the small sample size for other racial/ethnic groups, the current study focused solely on comparisons between non-Hispanic Black and non-Hispanic White participants. Fourth, the low prevalence of dementia prevented us from investigating its early-life contributors. Fifth, our model may suffer from overfitting due to the inclusion of a large set of early-life circumstances. Finally, while the exposures preceded the cognitive outcomes in this longitudinal study, the associations should not be interpreted as causal. To inform policies and interventions, additional research is needed to identify potential causal channels, for example, if the effects are directly driven by early-life environments or through parental exposure to unhealthy lifestyles and other unmeasured factors.

## Conclusions

This study identified early-life circumstances, especially educational factors, that may contribute to racial disparities in cognitive outcomes among older Americans. To slow cognitive decline and address racial disparities, additional research is needed to elucidate the mechanisms and inform the development of early-life interventions.

## Data Availability

All data produced in the present study are available upon reasonable request to the authors.

## Acknowledgements

This work was funded by the National Institute on Aging (R01AG077529; K01AG053408); Claude D. Pepper Older Americans Independence Center at Yale School of Medicine (P30AG021342); Yale Alzheimer’s Disease Research Center (P30AG066508); James Tobin Research Fund at Yale Economics Department; and Yale Macmillan Center Faculty Research Award. The funders had no role in the study design; data collection, analysis, or interpretation; in the writing of the report; or in the decision to submit the article for publication. The authors acknowledge helpful comments by participants and discussants at the various conferences, seminars, and workshops.

## Supplementary Online Content

## eAppendix A. Data Source

### Data Source 1. The HRS Core Survey

The Health and Retirement Study (HRS) has biennial interviews for collecting a wide range of information, including economics, health, marital, family status, and public and private support systems since 1992.^1^ Although the HRS has grown with the addition of new cohorts, the contents of the core survey have remained mostly consistent. In particular, the HRS core survey generally included multiple sections such as demographics and background, health, cognition, family structure and transfers, functional limitations, housing, physical measures, employment and pensions, disability, health services and insurance, expectations, assets and income, assets change, widowhood and divorce, and insurance. Additionally, the HRS has some experimental modules on specialized topics as part of the core survey. These modules only target a random subsample at the end of the core survey.^1,2^

In this study, variables of early-life demographics and socioeconomic status (SES) were constructed using the measures from the HRS core survey and/or related modules. Many of the variables were assembled from the RAND HRS files since RAND HRS groups have created user-friendly files with cleaned and processed variables with consistent and intuitive naming conventions, as well as model-based imputations.^3^ Otherwise, we assembled variables from the original HRS released core data from 1995 (when the variables were available) to 2018.

### Data Source 2. The Life History Mail Survey

The HRS Life History Mail Survey (LHMS) contains additional questions about respondents’ residential history, educational history, and other important early-life and family events. The 2015, 2017 Spring, and 2017 Fall versions were conducted in subsamples of HRS participants. The target subsample for the 2015 wave included all living HRS participants who were not included in the 2015 Consumption and Activities Mail Survey (CAMS) and who completed their most recent HRS core survey interview in English (rather than Spanish). The 2017 Spring wave included participants who were 2015 CAMS Sample Members who were still alive in 2017 and whose household was considered finalized on their 2016 core interview(s) by early March 2017 (members of finalized households either had completed core interviews or were considered final refusals for the core that wave). Lastly, the 2017 Fall sample included participants who were not included in the 2017 CAMS sample, and who did not return a 2015 LHMS questionnaire. The response rates and total enrolled participants for the 2015, 2017 Spring, and 2017 Fall waves respectively were 58% and 6,481 participants, 74% and 3,844 participants, and 28% and 1,444 participants.^4–6^ We assembled a range of variables for early-life circumstances surveyed in all three waves of the LHMS.

### Data Source 3. The Enhanced Face-to-Face Interview

In 2006, HRS initiated the Enhanced Face-to-Face (EFTF) interview with a mixed-mode design during the follow-up period in which a random half of HRS participants were assigned a face-to-face interview with physical and biological measures (e.g., salivary DNA samples) and a psychosocial questionnaire. The other half completed only the Core survey but were selected for the next (i.e., 2008) EFTF interview. A similar method was applied to the subsequent HRS survey. Several early-life circumstance variables in the present study related to trauma were assembled from this psychosocial questionnaire from 2006 to 2012.

In addition, polygenic risk scores (PGS) for a variety of phenotypes from respondents who provided salivary DNA between 2006 and 2012 were included in this study. Genotyping was conducted by the Center for Inherited Disease Research (CIDR) in 2011, 2012, and 2015, and principal component analysis was performed to identify population group outliers and to provide sample eigenvectors for association testing to adjust for potential population stratification. The final European American sample included all participants who self-reported as non-Hispanic White that had PC loadings within ± one standard deviations of the mean for eigenvectors 1 and 2 in the PC analysis of all unrelated study subjects. The final African American sample included all self-reported non-Hispanic Black Americans within two standard deviations of the mean of all self-identified Black Americans for eigenvector 1 and ± one standard deviation of the mean for eigenvector 2 in the PC analysis of all unrelated study subjects.

## eAppendix B. Early-Life Circumstances and Genetic Factors

Early-life circumstances involve all exposures during the early stages of life that may matter to long-term health outcomes. This study classified all early-life circumstances into the domains of cohort, regional, financial, health, trauma, family relationships, educational attainment, as well as educational quality and experiences. These domains were defined as a group of exposures. In addition, genetics were controlled in the analysis to capture individuals’ biological susceptibilities. Detailed variable definitions and constructions are presented below.

### Early-Life Cohort and Regional Factors

In this study, early-life demographics had two components, i.e., cohort and regional factors. Cohort factors measure whether the respondent was born in a special time period that may have profound impact on their prenatal and postnatal exposures, including birth during the Great Depression (1929-1933), birth during World War II (1941-1945). Regional factors measure respondents’ early-life aggregate exposure to regional contextual factors, including birth in the U.S. South, birth outside of the U.S., living in the U.S. South at age 10, and living outside of the U.S. at age 10. In HRS public data files, respondents’ birthplace and residence place at age 10 were reported at the census region level, including New England, Mid Atlantic, East North Central, West North Central, South Atlantic, East South Central, West South Central, Mountain, Pacific, other US/NA division, and not in US/US territory. Following existing literature, we defined participants’ birthplace (and childhood residence place) based on whether they were born (lived at age 10) in the South (including South Atlantic, East South Central, West South Central) or not. ^7^

### Early-life Financial Factors

We included several relevant variables that have been used before to measure respondents’ early-life financial status,^8–10^ such as whether the respondent relocated due to financial difficulties before age 16, whether their family had received financial assistance before respondents’ age 16. One additional variable—“father lost job”, indicating that the participants’ fathers experienced a significant unemployment spell (“several months or more”)—was included, as it was suggested to appropriately reflect early-life SES.^11^

### Early-life Health Events

To assess early-life health, we included 16 important health events occurring prior to age 18 from the Core, such as disabled for six months and severe head injury.^9,13^ Each variable for childhood health was coded as a dichotomous variable (0=no; 1=yes). All health outcomes which we included are as follows:

- Disabled for six months or more prior to age 18
- Injury to head which resulted in the loss of consciousness prior to age 18
- High blood pressure prior to age 18
- Heart issues prior to age 18
- Respiratory issues prior to age 18
- Diabetes prior to age 18
- Epilepsy prior to age 18
- Problems with speech prior to age 18
- Problems with hearing prior to age 18
- Problems with vision prior to age 18
- Depressed prior to age 18
- Other emotional or psychological disorder prior to age 18
- Learning disability prior to age 18
- Usage of drugs or alcohol prior to age 18
- Smoking cigarettes prior to age 18
- Parents smoked cigarettes prior to age 18

Respondents who had at least one of the aforementioned health event were coded as 1 and 0 otherwise.

### Early-life Traumas

We assembled several indicators of childhood traumatized experiences from the EFTF and the LHMS, which have been used before.^12^ Each variable for childhood trauma was coded as a dichotomous variable based on the answer (0=no; 1=yes). Since these trauma variables were from two different data sources (i.e., the EFTF interview as well as the 2015 Fall, 2017 Spring, and 2017 Fall LHMS), there was a slight difference in age frame in the questions (18 years old vs. age 16); hence we constructed two dichotomous trauma variables based on measures from each respective source. Specifically, respondents who had at least one of the trauma events from the EFTF were coded as 1 and 0 otherwise. Similarly, respondents with at least one of the trauma events from the LHMS were coded as 1 and 0 otherwise. The trauma events included in our study from each data source are provided below:

#### Traumas included from the EFTF

- Physical abused prior to age 18
- Parents used drugs or alcohol which caused problems prior to age 18
- Trouble with police prior to age 18
- Repeated school prior to age 18

#### Traumas included from the LHMS

- Spent any amount of time in an orphanage prior to age 16
- Spent any amount of time in a foster home prior to age 16
- One or more parents died prior to age 16
- Parents divorced prior to age 16
- Separated from mother for more than 6 months prior to age 16
- Separated from father for more than 6 months prior to age 16

### Early-Life Family Relationships

Family relationships included respondents’ self-rated quality of their relationship with their parents during childhood. Specifically, the relationship measures were coded as dichotomous variables (0=no; 1=yes) to indicate whether the respondent had good relationship with parents or not before age 18. The relationships were assessed respectively for father and mother.

### Early-Life Educational Factors

Educational factors consisted of two important components, including educational attainment (in years) and educational quality and experience.^14–16^ Educational attainment is assessed by years of educational attainment, which is a widely used early-life educational factor. Educational quality and experience included family education and quality of schooling and experience, which were assessed by a rich set of measures of education history that were uniquely collected in the HRS LHMS. To minimize the potential impact of recall bias, we defined dichotomous variables carefully to differentiate those with or without particular experience/exposure. We did not classify variables based on the degree of experience/exposure because it might suffer more from recall bias and could be less accurate due to ambiguity of the questions and answers.^17^ The classifications hence were defined based on the answer choices such as “not at all”, “none”, which can be clearly differentiated with others. Below we summarized the definition and construction of each factor within educational quality and experience.

**Family education** included:

- <u>Years of father’s educational attainment:</u> respondents’ reported father’s educational attainment (in years)
- <u>Years of mother’s educational attainment:</u> respondents’ reported mother’s educational attainment (in years)
- <u>Owned at least one shelf of books at age 10</u>: respondents were asked “When you were 10 years old, approximately how many books were in the place you lived (not counting magazines, newspapers, or your school books): none, enough to fill one shelf (11-25 books), enough to fill one bookcase (26-100 books), enough to fill two bookcase (101-200 books), or enough to fill more than two bookcase (more than 200 books).” Those who reported to have at least one shelf of books were coded as 1, and 0 otherwise.
- <u>Mother’s spent full time with children</u>: respondents were asked “What portion of the time did your mother work outside the home when you were growing up: all of the time, some of the time, or not at all?”. Those who answered “not at all” were coded as 1, and 0 otherwise.
- <u>Absence of time/attention from mother when needed</u>: respondents were asked “How much time and attention did your mother give you when you needed it (before you were 18 years old): a lot, some, a little, or not at all?” Those who answered “not at all” were considered to be absence of time/attention from mother and were coded as 1, and 0 otherwise.
- <u>Absence of effort in upbringing from mother</u>: respondents were asked “How much effort did your mother put into watching over you and making sure you had a good upbringing (before you were 18 years old): a lot, some, a little, or not at all?” Those who answered “not at all” were considered to be absence of effort from mother and were coded as 1, and 0 otherwise.
- <u>Absence of teaching about life from mother</u>: respondents were asked “How much did you mother teach you about life (before you were 18 years old): a lot, some, a little, or not at all?” Those who answered “not at all” were considered to be absence of teaching about life from mother and were coded as 1, and 0 otherwise.

For simplicity, in the main decomposition results, we reported the joint contribution of the latter four factors: 1) Mother spent full time with children before respondent’s age 18; 2) Absence of time/attention from mother when needed; 3) Absence of effort in upbringing from mother; 4) Absence of teaching about life from mother, and defined the domain as “Attention/Effort/Teaching from Mother”. We reported their contribution jointly because they all reflected similar attribute and the estimates would be easier to present and interpret than reporting the estimates individually. The individual estimates are also available upon request.

**Quality of schooling and experience** was assessed by respondents’ education history, which included:

- <u>Attended all minority school before college</u>: respondents were asked to list all of the primary, elementary, middle, junior high and high schools they attended; and for each school, they were asked whether “most children in the school were White, Black, Hispanic or others”. The school was considered to be a minority school if most children in the school were Black, Hispanic or others. If all the schools the respondents attended before college were minority schools, they were coded as 1 (and 0 otherwise).
- <u>Attended all public school before college</u>: respondents were asked to list all of the primary, elementary, middle, junior high and high schools they attended; and for each school, they were asked “Was this a public or private/religious school?”. If all the schools the respondents attended before college were public schools, they were coded as 1 (and 0 otherwise).
- <u>Preschool attendance</u>: respondents were asked “Did you attend a pre-school, nursery school, or other program before primary/elementary school?”. Those who answered “yes” were coded as 1, and 0 otherwise.
- <u>Learning problems in primary school</u>: respondents were asked “In primary or elementary school, did any teachers, principals or psychologists tell you or your parents that you had a problem with learning any of the usual school subjects below, including reading, writing, mathematics/arithmetic, and speaking/language?”, and they were asked “In primary or elementary school, were you or your parents ever told by a professional that you had any of the following problems, including Mental or emotional problems, dyslexia, attention deficit hyperactivity disorder (ADHD), and other learning disorders?”. Those who had any of the aforementioned learning problems in primary school were coded as 1, and 0 otherwise.
- <u>Learned foreign language in high school</u>: respondents were asked “Did you study a foreign language in high school?”. Those who answered “yes” were coded as 1, and 0 otherwise. Respondents who did not attend high school were coded as 0.
- <u>Learned creative arts in high school</u>: respondents were asked “In high school, did you take classes or spend time to do the following: 1) learn to play a musical instrument; 2) take singing lessons or sing in a chorus or choir; 3) learn woodwork or carpentry; 4) learn a craft (e.g., knitting, quilting, embroidery); 5) learn ballet or dance; and 6) learn to paint or draw or other art”. Those who learned any of the aforementioned creative arts in high school were coded as 1, and 0 otherwise. Respondents who did not attend high school were coded as 0.

### Genetic Factors

Genetic factors were assessed through the use of PGS, which is a score constructed based on variation in multiple genetic loci and their associated weights. It serves as the best prediction for the trait that can be made when accounting for variation in multiple genetic variants. All phenotypes for the four domains for genetic factors are listed below, and more information can be found in the document from HRS.^10^ Potential population stratification were accounted for.^18^

| **PGS Domains** | **Traits/phenotypes** |
| --- | --- |
| Anthropometric | Height |
|  | Body mass index |
|  | Waist circumference |
|  | Waist-to-Hip ratio |
| Disease/cognition/longevity | Coronary artery disease |
|  | Myocardial infarction |
|  | Type II diabetes |
|  | Alzheimer's disease IGAP 2013 with APOE |
|  | Alzheimer's disease IGAP 2019 with APOE |
|  | General cognition 2015 |
|  | General cognition 2018 |
|  | Longevity |
| Mental health/personality | Attention deficit/hyperactivity disorder PGC 2010 |
|  | Attention deficit/hyperactivity disorder PGC 2017 |
|  | Autism |
|  | Bipolar disorder |
|  | Mental health cross disorder |
|  | Major depressive disorder PGC 2010 |
|  | Major depressive disorder PGC 2018 |
|  | Subjective wellbeing |
|  | Neuroticism |
|  | Depressive symptoms |
|  | Schizophrenia |
|  | Extraversion |
| Smoking | Smoking initiation (ever/never) |
|  | Number of cigarettes smoked per day TAG 2010 |
|  | Number of cigarettes smoked per day GSCAN 2019 |

### Variable Missingness and Imputation

The table below summarizes the N (%) of missingness for outcomes, covariates, and early-life circumstances. Since we required participants to have at least one wave of valid cognitive assessment between 1995 to 2018, participants had no missing data regarding cognitive outcomes. In the Life History sample (N=9015), covariates had either no or limited missingness (<0.2%); and for 29 early-life circumstances included, 23 factors had 0-5% missingness, 5 factors had 5-10% missingness, and only 1 factor (years of father’s education) had slightly above 10% missingness. Similar patterns of missingness were observed in Life History sample who also had PGS data (N=7513).

Considering the large number of early-life circumstances included in the analysis, we performed multiple imputation to address item-level missingness of early-life factors.^19^ We followed a multivariate imputation procedure adopted in the HRS to impute the early-life factors, which is a sequential regression approach (i.e., chained equations) that create imputations through a sequence of multiple regressions.^20,21^ The imputation models included all the early-life factors (except genetic factors) and covariates included in the decomposition analyses, as well as other non-changing baseline demographics (e.g., birth year).^20,21^ Following existing guidelines, 20 imputed datasets were produced and analyzed; and the parameter estimates for each imputed datasets were pooled/combined to obtain the final decomposition results.^22,23^ Genetic factors were not imputed.

|  | Missingness in the Life History Sample (N=9015) |
| --- | --- |
| **Outcomes and Covariates** |  |
| Cognitive score | 0 (0.00%) |
| Cognitive impairment prevalence | 0 (0.00%) |
| Age | 0 (0.00%) |
| Sex | 0 (0.00%) |
| Married/cohabiting | 12 (0.13%) |
| **Early-Life Circumstances** |  |
| Born during World War II (1941-1945) | 0 (0.00%) |
| Born during the Great Depression (1929-1933) | 0 (0.00%) |
| Born in the South | 2 (0.02%) |
| Born outside of the US | 2 (0.02%) |
| Lived in the South at age 10 | 1 (0.01%) |
| Lived outside of the US at age 10 | 1 (0.01%) |
| Relocated due to financial difficulties before age 16 | 39 (0.43%) |
| Family received financial help before age 16 | 159 (1.8%) |
| Father unemployed before respondent's age 16 | 704 (7.8%) |
| Household size at age 10, median (IQR) | 95 (1.1%) |
| Adverse childhood health events | 103 (1.1%) |
| Childhood trauma events (EFTF) | 29 (0.32%) |
| Childhood trauma events (LHMS) | 127 (1.4%) |
| Good relationship with mother before age 18 | 446 (4.9%) |
| Good relationship with father before age 18 | 712 (7.9%) |
| Years of educational attainment | 38 (0.42%) |
| Years of father's educational attainment | 1174 (13.0%) |
| Years of mother's educational attainment | 666 (7.4%) |
| Owned at least one shelf of books at age 10 | 193 (2.1%) |
| Mother spent full time with children before age 18 | 171 (1.9%) |
| Absence of time/attention from mother when needed | 282 (3.1%) |
| Absence of effort in upbringing from mother | 288 (3.2%) |
| Absence of teaching about life from mother | 289 (3.2%) |
| Attended all minority school before college | 863 (9.6%) |
| Attended all public school before college | 632 (7.0%) |
| Preschool attendance | 238 (2.6%) |
| Learning problems in primary school | 105 (1.2%) |
| Learned foreign language in high school | 339 (3.8%) |
| Learned creative arts in high school | 313 (3.5%) |

## eAppendix C. Decomposition Analysis

***Blinder-Oaxaca Decomposition (BOD)*** was used to measure the extent to which early-life circumstances may individually and collectively contribute to racial disparities in cognitive outcomes, including cognitive score and cognitive impairment, between *White* and *Black* participants. The BOD decomposes mean differences in regression outcomes in a counterfactual manner, and is widely used in economics to understand racial disparities.^24–34^ It allows us to separate cognitive differences between White and Black participants into a part that is explained by differences in early-life circumstances, versus a part that cannot be accounted for by such differences.^26,35^ We tested the hypothesis that early-life circumstances contribute significantly and sizably to racial disparities in cognitive outcomes.

We focused on comparing racial disparities in cognitive outcomes between White and Black participants, and examined the extent to which racial differences were attributed to the differences in early-life circumstances.^36,37^ Formally, the differences in cognitive outcomes between White (*W*) and Black (*B*) participants can be formulated as,

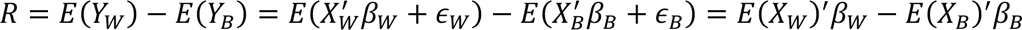

where *E*(*Y*_*l*_) denotes the expected value of cognitive outcomes for race *l*, *l* ∈ (*W*, *B*); and *X*_*l*_is a matrix that contains early life circumstances and constants. The second and the third equalities hold under the assumptions of linear model, 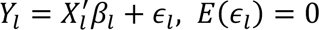. The equation can be decomposed as follows.

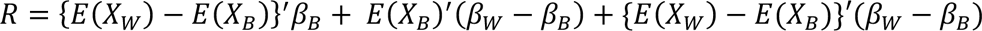

The first component {*E*(*X_W_*) − *E*(*X_B_*)}′*β_B_* represents the part of the differential that is attributable to group differences in early-life circumstances. The second component *E*(*X_B_*)′(*β_W_* – *β_B_*) measures the contribution of differences in coefficients. The third component {*E*(*X_W_*) − *E*(*X_B_*)}′(*β_W_* – *β_B_*) is an interaction term accounting for both the differences in early-life circumstances and coefficients. Our parameter of interest is the first component, which measures the expected changes in Black participants’ mean cognition if they had the same levels of early life circumstances as White participants (i.e., overall contribution of early-life circumstances). To understand how much of the racial disparities in cognitive outcomes are accounted for by individual characteristics, this component can be further decomposed as,

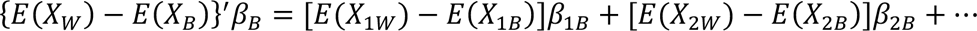

where *E*(*X*_1_), *E*(*X*_2_), … are the expectation of individual characteristics; and *β*_1_, *β*_2_, … are the associated coefficients. [*E*(*X*_1*W*_) − *E*(*X*_1*B*_)]*β*_1*B*_, thus captures the individual contribution of the differences in *X*_1_.

In this study, the decompositions were conducted at both the overall and the variable level to evaluate the collective and individual contribution of early-life circumstances. For continuous outcome (i.e., cognitive score), BOD was conducted using a linear decomposition method, while for dichotomous outcome (i.e., cognitive impairment), a nonlinear decomposition method was employed.^24–34,38^

---

**eTable 1.**
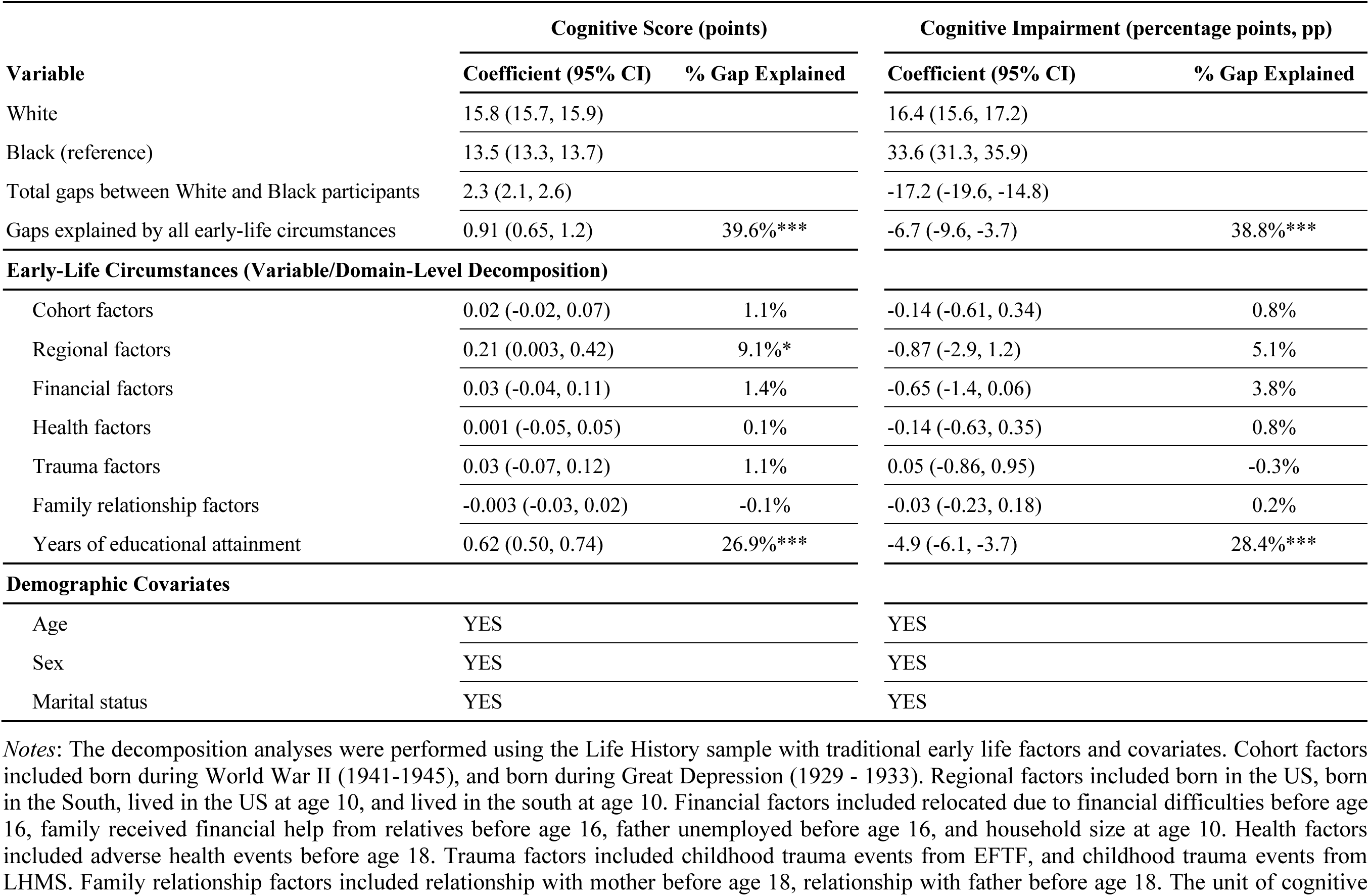
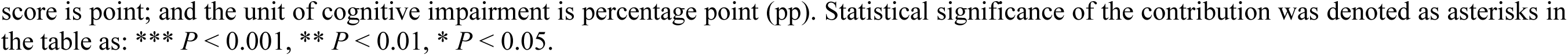
Numerical Estimates of the Early-Life Contribution to Racial Gaps in Cognition, LifeHistory**_1_** (N=9015)

**eTable 2.** Numerical Estimates of the Early-Life Contribution to Racial Gaps in Cognition, LifeHistory**_2_** (N=9015)

| Variable | Cognitive Score (points) |  | Cognitive Impairment (percentage points, pp) |  |
| --- | --- | --- | --- | --- |
|  | Coefficient (95% CI) | % Gap Explained | Coefficient (95% CI) | % Gap Explained |
| White | 15.8 (15.7, 15.9) |  | 16.4 (15.6, 17.2) |  |
| Black (reference) | 13.5 (13.3, 13.7) |  | 33.6 (31.3, 35.8) |  |
| Total gaps between White and Black participants | 2.3 (2.1, 2.6) |  | -17.2 (-19.6, -14.8) |  |
| Gaps explained by all early-life circumstances | 1.5 (1.1, 2.0) | 65.9%*** | -14.6 (-19.0, -10.3) | 85.1%*** |
| **Early-Life Circumstances (Variable/Domain-Level Decomposition)** |  |  |  |  |
| Traditional Early-Life Factors (Grouped) | 0.73 (0.46, 1.0) | 31.8%*** | -5.8 (-8.4, -3.2) | 33.8%*** |
| Cohort factors | 0.03 (-0.02, 0.08) | 1.2% | -0.18 (-0.67, 0.31) | 1.0% |
| Regional factors | 0.15 (-0.06, 0.37) | 6.7% | -0.72 (-2.9, 1.5) | 4.2% |
| Financial factors | 0.01 (-0.07, 0.09) | 0.5% | -0.54 (-1.26, 0.18) | 3.1% |
| Health factors | -0.001 (-0.05, 0.05) | -0.1% | -0.10 (-0.61, 0.41) | 0.6% |
| Trauma factors | 0.02 (-0.08, 0.11) | 0.7% | 0.11 (-0.85, 1.07) | -0.6% |
| Family relationship factors | 0.0004 (-0.02, 0.02) | 0.0% | -0.07 (-0.30, 0.15) | 0.4% |
| Years of educational attainment | 0.52 (0.41, 0.64) | 22.7%*** | -4.3 (-5.5, -3.1) | 25.1%*** |
| Additional Early-Life Educational Factors (Grouped) | 0.79 (0.38, 1.19) | 34.1%*** | -8.8 (-13.1, -4.6) | 51.3%*** |
| Years of father's educational attainment | -0.01 (-0.15, 0.13) | -0.4% | 0.27 (-1.0, 1.6) | -1.6% |
| Years of mother's educational attainment | 0.02 (-0.09, 0.14) | 1.0% | -0.31 (-1.4, 0.83) | 1.8% |
| Owned books at age 10 | -0.01 (-0.08, 0.06) | -0.5% | 0.60 (-0.08, 1.27) | -3.5% |
| Attention/effort/teaching from mother | -0.01 (-0.13, 0.11) | -0.6% | -0.29 (-1.4, 0.84) | 1.7% |
| Attended all minority school before college | 0.62 (0.24, 0.99) | 26.7%** | -7.0 (-11.0, -2.9) | 40.6%*** |
| Attended all public school before college | 0.02 (-0.07, 0.12) | 0.9% | -0.55 (-1.6, 0.49) | 3.2% |
| Preschool attendance | 0.02 (-0.01, 0.05) | 0.9% | -0.26 (-0.59, 0.08) | 1.5% |
| Learning problems in primary school | 0.06 (0.02, 0.10) | 2.7%** | -0.63 (-1.0, -0.26) | 3.7%*** |
| Learned foreign language in high school | 0.06 (0.02, 0.11) | 2.8%** | -0.58 (-1.0, -0.14) | 3.4%** |
| Learned creative arts in high school | 0.01 (-0.01, 0.03) | 0.5% | -0.09 (-0.24, 0.06) | 0.5% |
| **Demographic Covariates** |  |  |  |  |
| Age | YES |  | YES |  |
| Sex | YES |  | YES |  |
| Marital status | YES |  | YES |  |

**eFigure 1.**
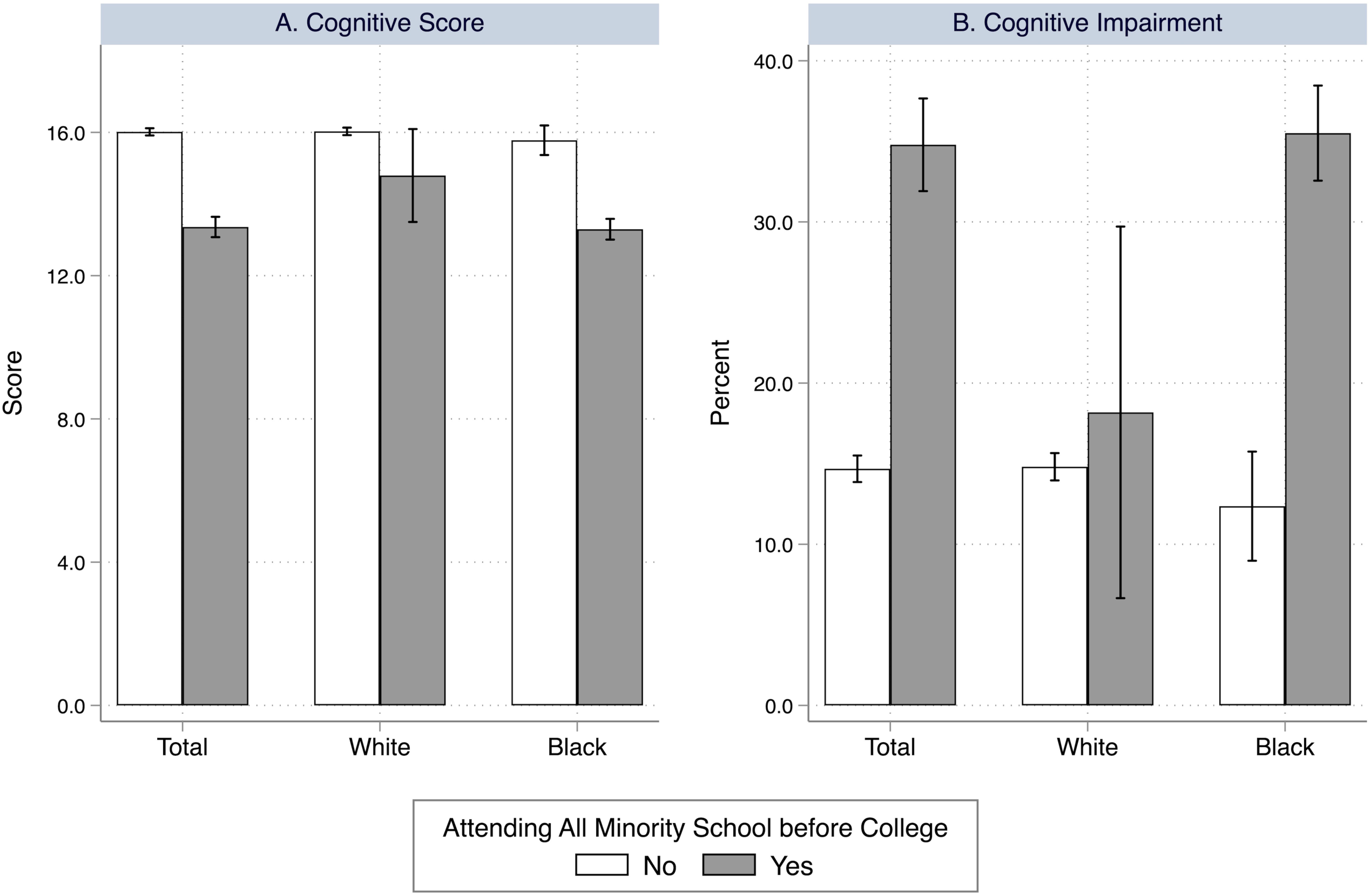
Differences in Cognitive Outcomes between Participants Who Attended All Minority Schools before College and Those Who Did Not in the Life History Sample. ***Notes*:** This figure presents the average cognitive outcomes for participants who attended all minority schools before college (in gray color) and those who did not (in white color). The estimates were obtained for the all participants (Total), White participants, and Black participants in the Life History sample. Panel A shows the average cognitive score; and Panel B shows the average proportion of cognitive impairment (%). The vertical bars represent the mean estimates, and the vertical lines present the 95% confidence interval. Sample sizes were respectively 7,097 (No) and 1055 (Yes) for Total participants, 6733 (No) and 44 (Yes) for White participants; and 364 (No), and 1011 (Yes) for Black participants.

**eFigure 2.**
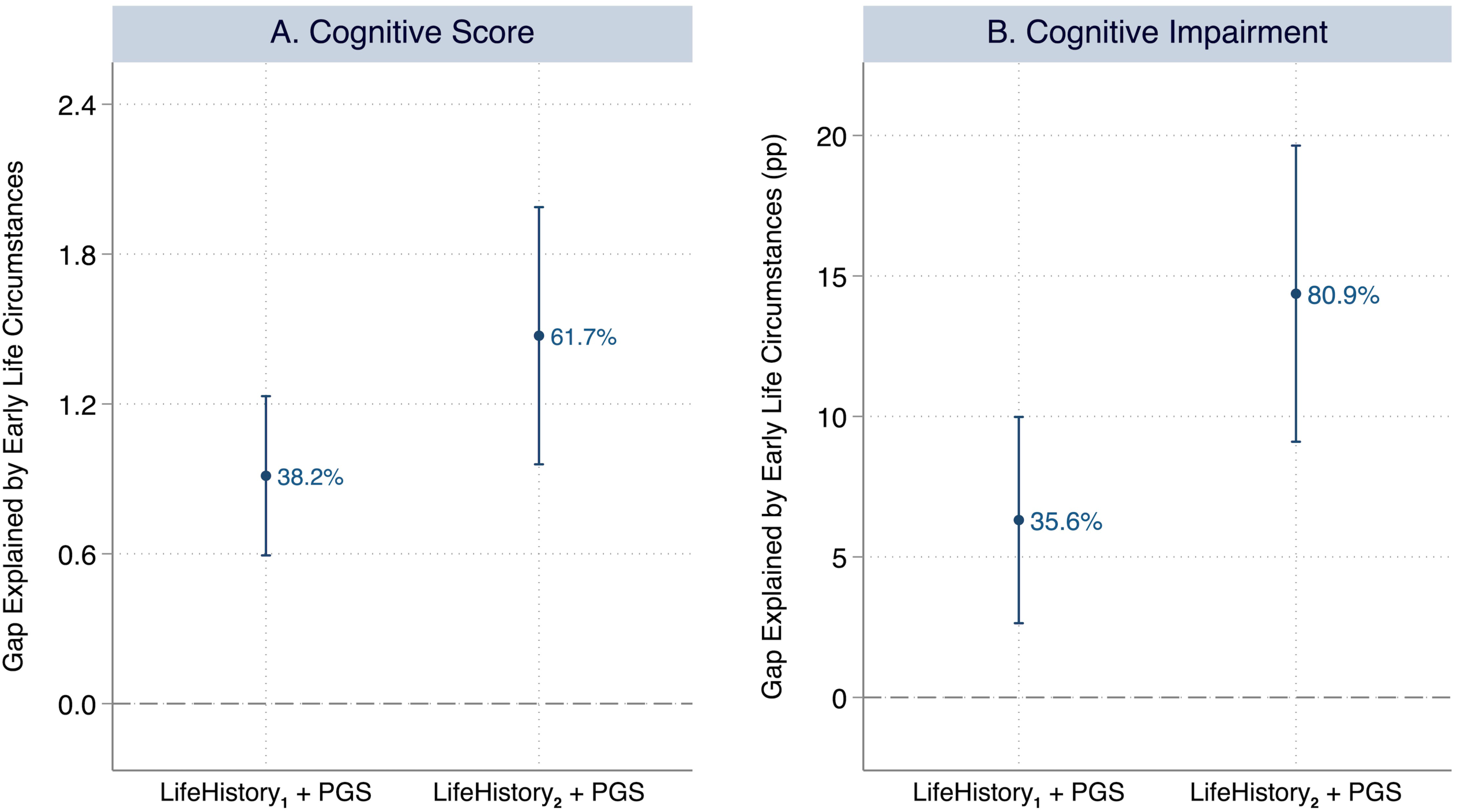
Absolute and Relative Contribution of Overall Early-Life Circumstances to Racial Gaps in Cognition between White and Black Participants with Genetic Adjustment (N=7513) ***Notes*:** Panel A presents the decomposition results for cognitive score, a continuous variable ranging from 0 to 27; and Panel B presents the decomposition results for cognitive impairment, a dichotomous variable indicating whether individuals were classified as cognitive impaired (0/1). For each cognitive outcome, X axis denotes the models being examined, including LifeHistory**_1_** (with genetic adjustment), LifeHistory**_2_** (with genetic adjustment). In LifeHistory_1_, traditional early-life factors were included to perform the decomposition. In LifeHistory_2_, early-life educational quality and experience were additionally added. All decompositions adjust for demographic and biological covariates including age, sex and marital status as well as genetic factors (i.e., PGS). In Panels A and B, Y axis denotes the racial gap in cognitive outcomes between White and Black participants that was explained by their differences in early-life circumstances, representing the absolute contribution of early-life circumstances to racial gaps in cognition. In Panel A, the unit of cognitive score is point; and in Panel B, the unit of cognitive impairment is percentage point (pp). The point estimates are plotted as circles and their 95% confidence interval are plotted as vertical lines. The relative contributions of the early-life circumstances (%) in each setting were also presented alongside the vertical lines, denoting the percentage of gaps in cognition that was explained by early-life circumstances. The pooled estimates were obtained using multiple imputation with 20 imputed datasets.

**eFigure 3.**
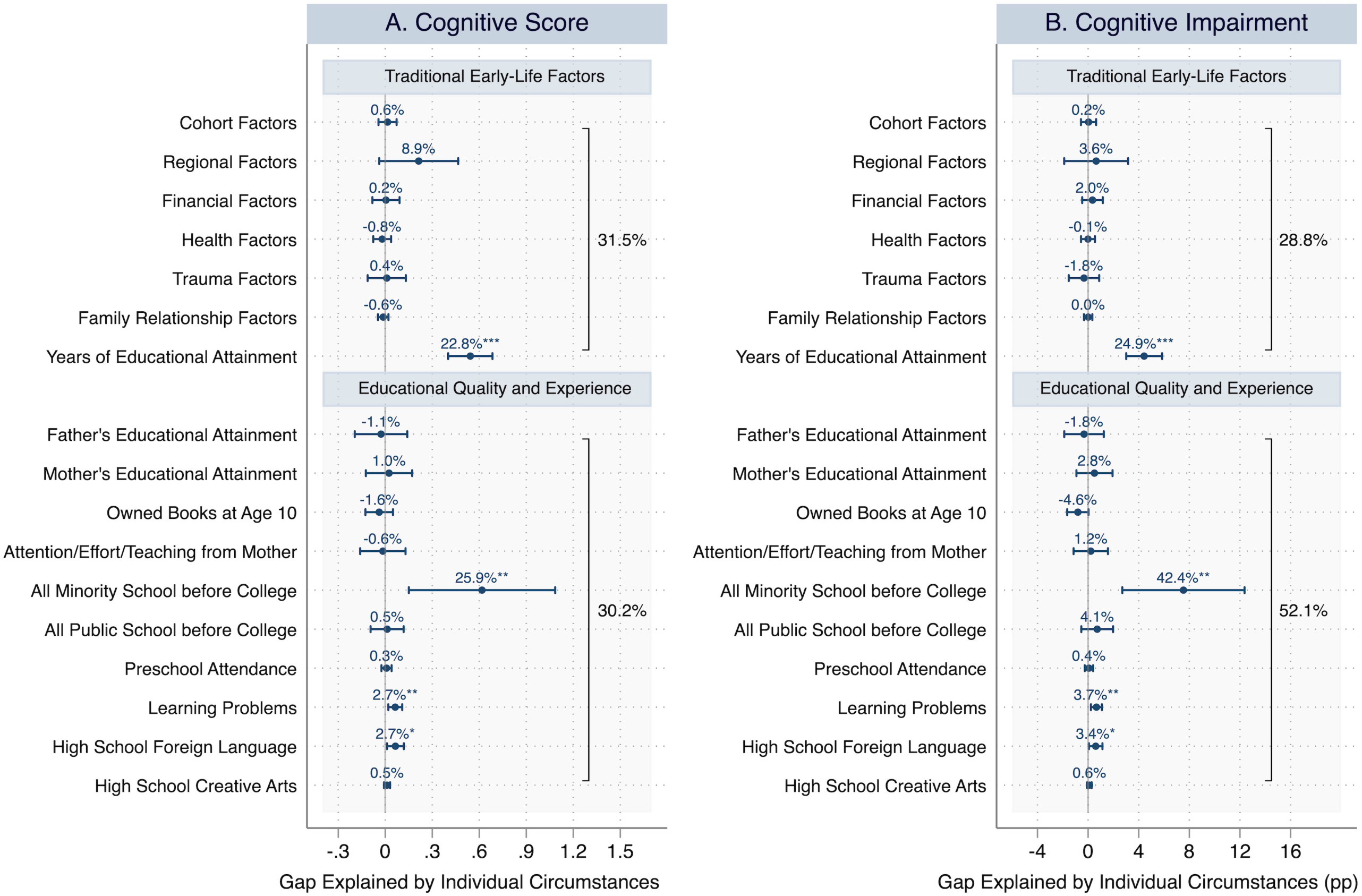
Absolute and Relative Contribution of Individual Early-Life Circumstances to Racial Gaps in Cognition between White and Black Participants with Genetic Adjustment (N=7513) ***Notes*:** Panel A shows the decomposition results for cognitive score, a continuous variable ranging from 0 to 27; Panel B shows the decomposition results for cognitive impairment, a dichotomous variable indicating whether individuals were classified as cognitive impaired (0/1), or not. For each cognitive outcome, Y axis denotes the variables or domains being examined; X axis denotes the amount of racial gap in cognition between White and Black participants that was explained by their differences in the individual early-life circumstances (i.e., absolute contribution). In Panel A, the unit of cognitive score is point; and in Panel B, the unit of cognitive impairment is percentage point (pp). The point estimates are plotted as circles and their 95% confidence interval are plotted as horizontal lines. The relative contributions of the early-life circumstances (%) in each setting were presented alongside the horizontal lines, denoting the percentage of gaps in cognition that was explained by early-life circumstances. The decompositions were performed using Life History sample with genetic factors, and the decompositions adjust for demographic and biological covariates including age, sex and marital status, as well as genetic factors (i.e., PGS). The pooled estimates were obtained using multiple imputation with 20 imputed datasets; and the numerical estimates are available upon request. Asterisks denote the statistical significance of the contribution: *** *P* < 0.001, ** *P* < 0.01, * *P* < 0.05.

